# Resting State Functional Connectivity is Decreased Globally Across the *C9orf72* Mutation Spectrum

**DOI:** 10.1101/2020.08.10.20171991

**Authors:** Rachel F Smallwood Shoukry, Michael G Clark, Mary Kay Floeter

**Affiliations:** Motor Neuron Disease Unit, National Institute of Neurological Disorders and Stroke, National Institutes of Health, Bethesda, MD, USA

**Keywords:** **C9orf72**, **amyotrophic lateral sclerosis**, **behavioral variant frontotemporal dementia**, **presymptomatic**, **resting state fMRI**, **graph theory**

## Abstract

A repeat expansion mutation in the *C9orf72* gene causes amyotrophic lateral sclerosis (ALS), frontotemporal dementia (FTD), or symptoms of both, and has been associated with gray and white matter changes in brain MRI scans. We used graph theory to examine the network properties of brain function at rest in a population of mixed-phenotype *C9orf72* mutation carriers (C9+). Twenty-five C9+ subjects (presymptomatic, or diagnosed with ALS, behavioral variant FTD (bvFTD), or both ALS and FTD) and twenty-six healthy controls underwent resting state fMRI. When comparing all C9+ subjects with healthy controls, both global and connection-specific decreases in resting state connectivity were observed, with no substantial reorganization of network hubs. However, when analyzing subgroups of the symptomatic C9+ patients, those with bvFTD (with and without comorbid ALS) show remarkable reorganization of hubs compared to patients with ALS alone (without bvFTD), indicating that subcortical regions become more connected in the network relative to other regions. Additionally, network connectivity measures of the right hippocampus and bilateral thalami increased with increasing scores on the Frontal Behavioral Inventory, indicative of worsening behavioral impairment. These results indicate that while *C9orf72* mutation carriers across the ALS-FTD spectrum have global decreased resting state brain connectivity, phenotype-specific effects can also be observed at more local network levels.

## 1. Introduction

A repeat expansion mutation in the *C9orf72* gene is the most frequent cause of familial amyotrophic lateral sclerosis (ALS) and familial frontotemporal dementia (FTD) in populations of Northern European origin (1, 2) accounting for 5-10% of sporadic cases of these disorders (3). Carriers of the *C9orf72* mutation (hereafter referred to as C9+) can present with clinical symptoms of ALS, FTD, or with combinations of motor, cognitive, and behavioral symptoms (4-7). Compared to patients with sporadic ALS or FTD, neuroimaging studies in C9+ ALS and FTD patients show more pronounced atrophy, particularly of subcortical structures and extramotor cortical regions (4, 8-15). Although subtle structural changes can be detected in groups of presymptomatic C9+ carriers (9, 16-18), most structural changes are found later in the disease course, when symptoms are manifest. In individual C9+ carriers, the structural changes may represent a hybrid pattern between those described for sporadic ALS and sporadic FTD, appearing to reflect the relative balance of motor and cognitive-behavioral dysfunction (10, 19-21).

Functional connectivity changes also occur in patients with ALS and FTD. In sporadic and C9+ FTD, intrinsic functional connectivity is reduced in the salience networks, frontal and temporal regions, and thalamic networks (13, 22-24). Findings from resting state fMRI studies in sporadic ALS are less consistent. Most reports find increased connectivity, particularly in the sensorimotor network and default mode network (25-28). However, others report decreased connectivity (29-32), or mixtures of increased and decreased network connectivity (33-36). We hypothesized that functional imaging in C9+ carriers would show a hybrid pattern on a continuum of those seen in ALS and FTD reflecting the relative balance of motor and cognitive-behavioral dysfunction in each patient. To examine this hypothesis, we evaluated changes in network measures and their association with clinical measures of motor and cognitive-behavioral dysfunction using graph theory metrics. In this analysis, each brain region is represented as a node and the relationship between two brain regions is represented as an edge connecting the two nodes (37). Graph theory allows us to quantify whole-brain network properties, as well as how regions interact with each other as part of a larger network.

We first compared differences in network measures of functional connectivity in a heterogeneous group of C9+ carriers to healthy controls to identify changes associated with the *C9orf72* mutation itself. We then compared C9+ carriers with ALS alone (C9+ FTD−) to C9+ patients with behavioral variant FTD (bvFTD) or ALS-FTD within the cohort. We hypothesized that C9+ with bvFTD/ALS-FTD would exhibit changes in networks associated with cognitive-behavioral function whereas C9+ FTD – ALS patients would exhibit changes in motor networks, and that network measures would correlate with clinical measures of motor or cognitive-behavioral function.

## 2. Methods

### 2.1 Participants

Twenty-five carriers of the *C9orf72* expansion mutation (**Table 1**) were recruited from across the United States though online advertising, organizational outreach, and physician referrals between 2013 and 2016. All subjects gave written informed consent in accordance with an IRB-approved protocol. Inclusion criteria required the C9+ subjects to have >30 repeats in the *C9orf72* gene as established by repeat prime polymerase chain reaction in a CLIA certified lab. They were not excluded for having other comorbid conditions. All C9+ subjects were examined by an experienced neurologist and underwent electromyography and cognitive testing to determine their clinical diagnosis as previously reported (10). ALS was diagnosed using the 2015 revised El Escorial criteria (38). The International Consensus Criteria for behavioral variant FTD (39) were used for diagnosis of possible, probable, or definite bvFTD. The cohort consisted of C9+ subjects classified as being presymptomatic (N=7), or as having ALS only (N=9), bvFTD only (N=3), or both ALS and bvFTD (N=6). C9+ patients were administered the revised ALS functional rating scale (ALSFRS-R; (40)) to quantify motor impairment related to ALS and the frontal behavioral inventory (FBI; (41)) to assess behavioral impairment related to bvFTD. Twenty-six healthy controls (HC) underwent the same imaging protocol as the C9+ patients as part of a separate IRB-approved study. All healthy controls had normal neurological examinations and a normal cognitive screening test.

**Table 1.** Demographic information of study groups and sub-groups.

|  | **N (males)** | **Age (years)** | **ALSFRS-R** | **FBI** |
| --- | --- | --- | --- | --- |
| **All C9+** | **25 (14)** | **51.74 ± 11.95** | **42.8 ± 6.7** | **0.16 ± 0.17** |
| ALS | 9 (4) | 51.85 ± 9.95 | 41.6 ± 3.8 | 0.08 ± 0.08 |
| ALS-FTD | 6 (6) | 60.72 ± 10.19 | 41.8 ± 5.3 | 0.31 ± 0.08 |
| bvFTD | 3 (3) | 60.59 ± 5.11 | 46.0 ± 2.7 | 0.36 ± 0.2 |
| **HC** | **26 (16)** | **52.33±8.79** | **-** | **-** |

### 2.2 Imaging protocol

Participants underwent MRI scanning on a GE 3T scanner. Two T1 FSPGR anatomical scans were collected (TI = 450 ms, a =13°, voxel size = 1×70.938×70.938 mm). Resting state functional scans were collected during which subjects were instructed to stay awake, keep their eyes open, and think random thoughts (TR=2000ms, TE=30ms, flip angle=77, voxel size = 3.75×73.75×73.8 mm, FOV = 64×764 cm, 40 slices, 214 volumes).

### 2.3 Image processing

Anatomical MRI data were processed in FreeSurfer and each subject’s gray matter was parcellated into 82 volumes of interest (VOIs). These VOIs consisted of 34 cortical regions per hemisphere based on the Desikan-Killiany atlas (42) plus 14 subcortical VOIs, excluding the cerebellum **(Table 2)**. Resting state functional data were preprocessed using FSL and custom MATLAB scripts. Preprocessing steps included motion and slice time correction, volume scrubbing based on motion outliers (identified by FSL’s calculation of framewise displacement and the derivative of the time-course of root mean square intensity across voxels (DVARS; (43)), regression of motion parameters and white matter and CSF signals, and bandpass filtering between 0.01-0.1 Hz. Each subject’s functional image was registered to their structural image. The anatomical parcellation and segmentation were applied to the functional time series to extract the 82 VOIs, and all the voxels within each VOI were averaged at every time point to create a time series of average signal for each VOI.

**Table 2.** –Volumes of interest used in the analysis and their abbreviations and color scheme as displayed in Figure 3. Each region had a right and left representation. Blue – frontal lobe; green – limbic system; yellow – basal ganglia; orange – temporal lobe; red – parietal lobe; purple – occipital lobe.

| Region name | Abbreviation | Region name | Abbreviation |
| --- | --- | --- | --- |
| Frontal pole | Front-pole | Caudate |  |
| Superior frontal gyrus | SFG | Putamen |  |
| Lateral orbitofrontal cortex | Lat-OFC | Accumbens area | Accumbens |
| Medial orbitofrontal cortex | Med-OFC | Pallidum |  |
| Rostral middle frontal gyrus | Rost-MFG | Temporal pole | Temp-pole |
| Pars triangularis | Pars-tri | Superior temporal gyrus | STG |
| Pars orbitalis | Pars-orb | Transverse temporal gyrus | Transverse |
| Pars opercularis | Pars-operc | Banks of the superior temporal sulcus | Banks-STS |
| Caudal middle frontal gyrus | Caud-MFG | Middle temporal gyrus | MTG |
| Precentral gyrus | Precentral | Inferior temporal gyrus | ITG |
| Paracentral lobule | Paracentral | Fusiform gyrus | Fusiform |
| Rostral anterior cingulate cortex | Rost-ACC | Insula |  |
| Caudal anterior cingulate cortex | Caud-ACC | Postcentral gyrus | Postcentral |
| Posterior cingulate cortex | PCC | Supramarginal gyrus | Supramarg |
| Isthmus of the cingulate cortex | Isth-CC | Superior parietal cortex | SPC |
| Amygdala |  | Inferior parietal cortex | IPC |
| Thalamus |  | Precuneus |  |
| Hippocampus |  | Cuneus |  |
| Entorhinal cortex | Entorhinal | Lateral occipital cortex | Lat-OC |
| Parahippocampal gyrus | Parahipp | Pericalcarine cortex | Pericalc |
|  |  | Lingual gyrus | Lingual |

### 2.4 Graph theory analysis and statistics

A connectivity matrix was formed with each row and column representing a node (VOI) and each cell representing an edge with strength equal to the Pearson correlation coefficient (R) of the row/column pair. Fisher transformation was applied to permit multiple linear regression of age and gender effects, followed by back transformation to R. The matrices were then thresholded by setting all connections below a specific R to zero. Thresholds from 0 to 0.7 were tested in increments of 0.1; thresholds >0.7 resulted in matrices too sparse for the calculation of may graph metrics. Thresholds between 0 to 0.4 yielded similar statistical results in global metrics, so R ≥ 0.2 was selected as the representative threshold for reporting. Graph theory metrics were calculated using custom MATLAB scripts and the Brain Connectivity Toolbox (https://sites.google.com/site/bctnet (37)).

Two group analyses were performed: 1) all C9+ versus HC and 2) a sub-analysis of C9+ subjects in which symptomatic bvFTD+ patients (bvFTD & ALS-FTD, n=9) were compared to symptomatic ALS-only patients (ALS only, n=9). Group differences for both analyses were evaluated using global, nodal, and edge metrics (**Table 3**). Global metrics included network density, mean connection strength, mean node clustering coefficient, mean node path length, and modularity score. Nodal measures included node strength, closeness centrality, betweenness centrality, within-module degree Z score, and participation coefficient. Hubs, or nodes that are particularly highly connected and involved within the network, were identified with a composite hub score that was calculated by summing the Z scores of the five nodal measures.

**Table 3.**
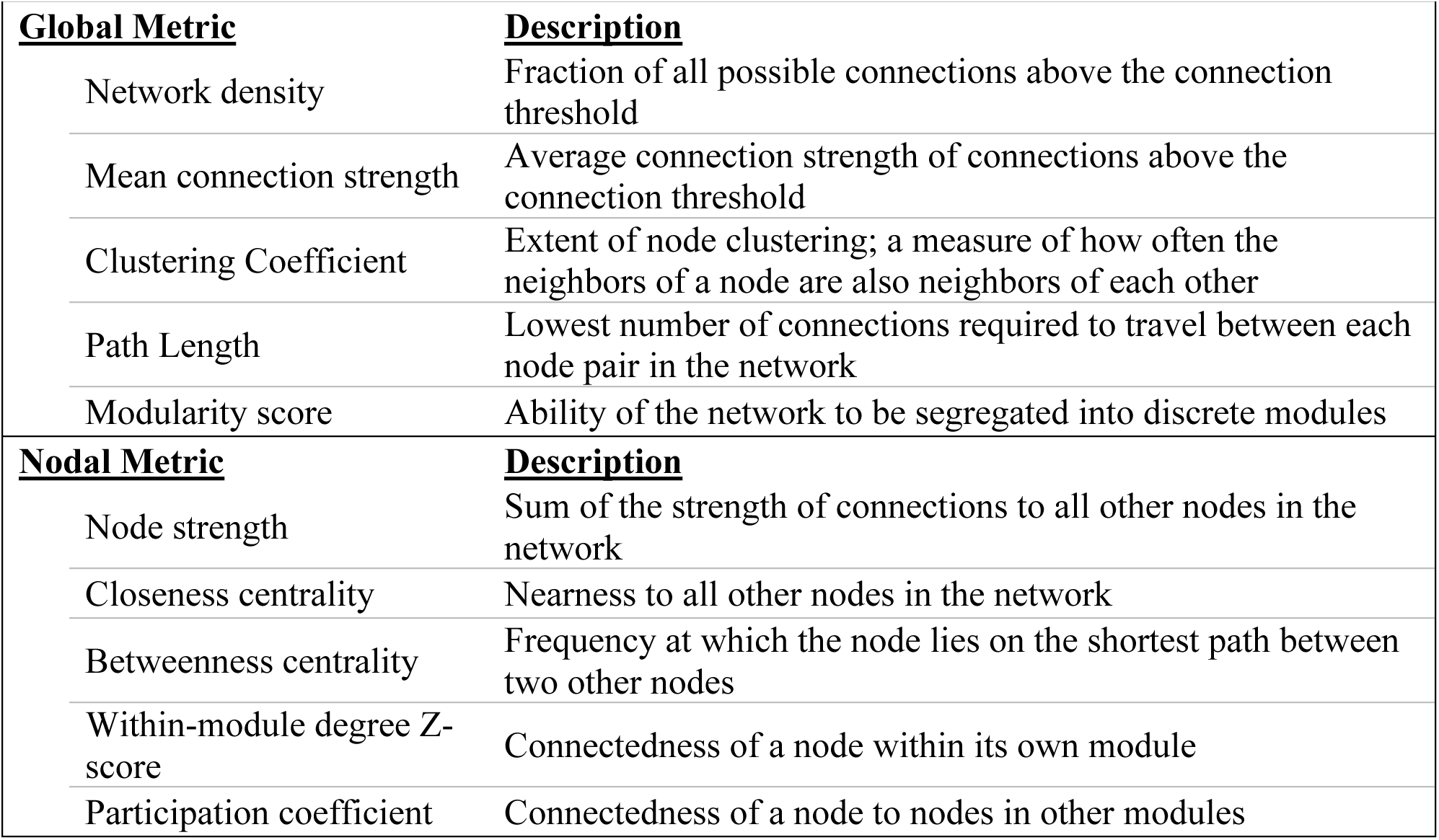
– Description of graph analysis metrics.

Permutation testing was used to calculate effective p values (44). In each permutation, subjects’ group assignments were randomly shuffled and the difference in the metric of interest between the two shuffled groups was calculated. The effective p value was defined as the fraction of total permutations in which the shuffled groups had a larger magnitude of difference in the graph metric than the actual study groups. 1×10^3^ permutations were used to ensure a sufficient number of significant figures for reporting.

## 3. Results

### 3.1 Comparison of C9+ carriers with Healthy Controls

#### 3.1.1 Global graph metrics – C9+ vs. HC

Analysis of global measures revealed that the C9+ carrier group had significantly lower global network density than HC, with fewer connections with strength greater than the correlation threshold (**Figures 1 & 2**). Those edges that survived thresholding had a significantly lower mean connection strength for C9+ than HC (**Figure 2**). The clustering coefficients were also significantly different between groups (p=0.018). However, because path length and modularity score are sensitive to network density (45), any significant differences are likely to be influenced by the different network densities, so they were not further explored.

**Figure 1.**
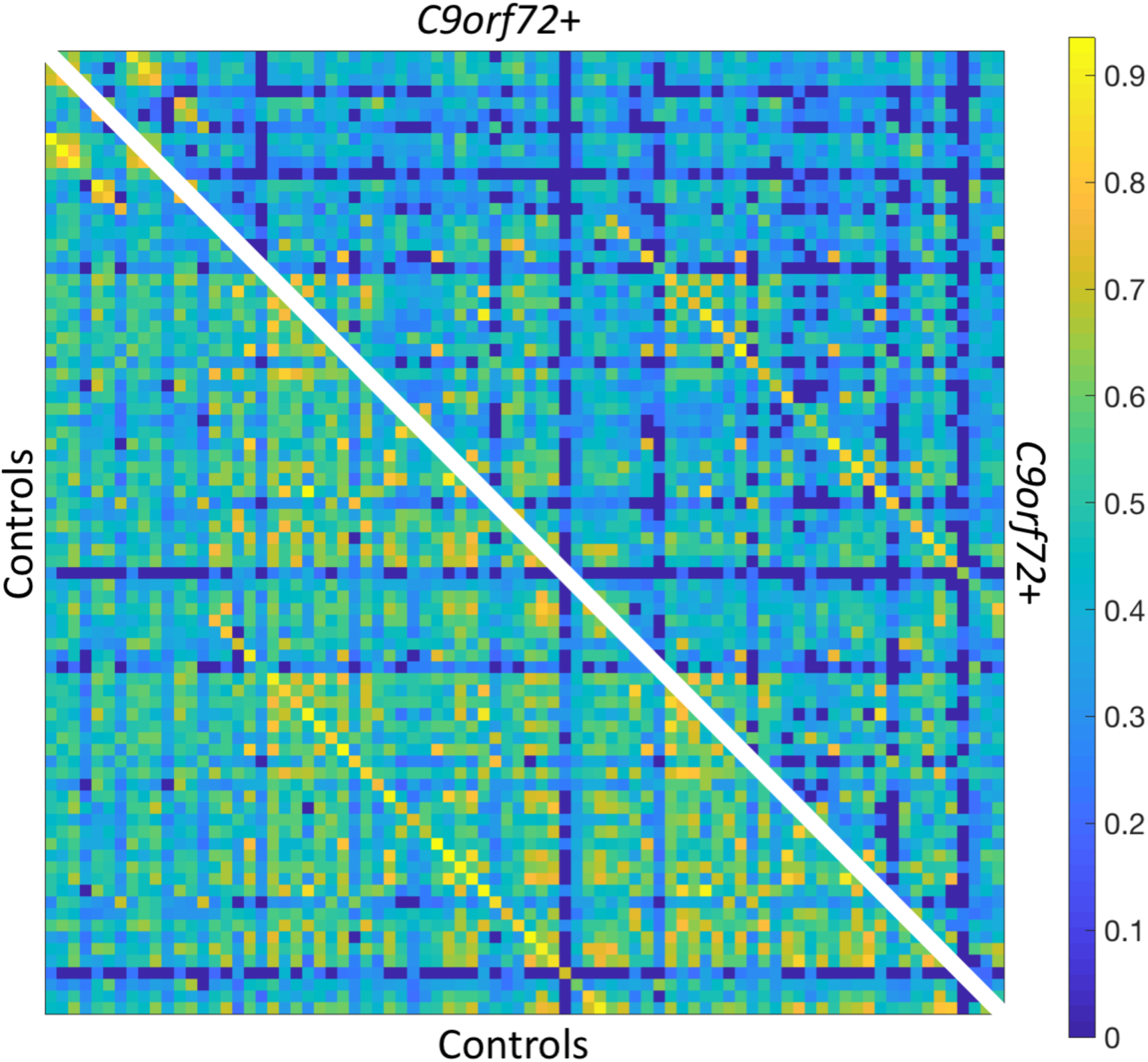
- Resting state functional connectivity matrix thresholded at R≥0.2. Bottom left: HC mean connectivity matrix. Top right: C9+ mean connectivity matrix. Scale bar shows Pearson correlation coefficient.

**Figure 2.**
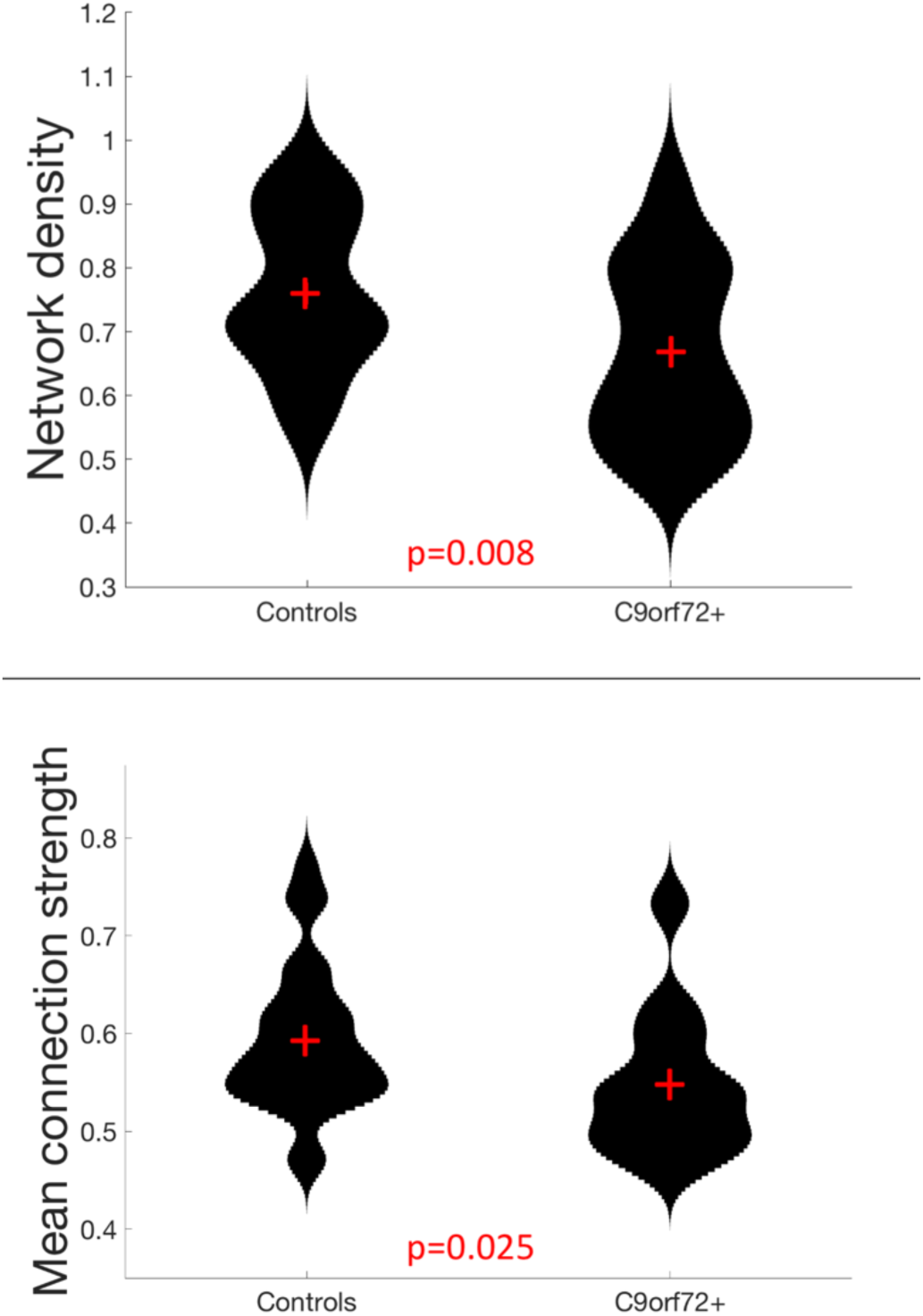
- Global network measures in which C9+ carriers significantly differed from HC.

#### 3.1.2 Network hubs – C9+ carriers vs. Healthy Controls

There was minimal reorganization of hub nodes in C9+ carriers compared to the healthy control group. The composite hub score showed that 75% of the nodes comprising the top 20% of hub scores were the same for HC and C9+ carriers (**Table 4**). Moreover, with the exception of one node in the top 20% in each group, hubs were ranked within the top 30% in the other group or were the contralateral pair to one of the top 20% nodes. The single hub that was highly ranked in healthy controls but not C9+ was the left lateral orbitofrontal cortex. The single hub that ranked highly in the C9+ carrier group but not healthy controls was the left thalamus.

**Table 4.**
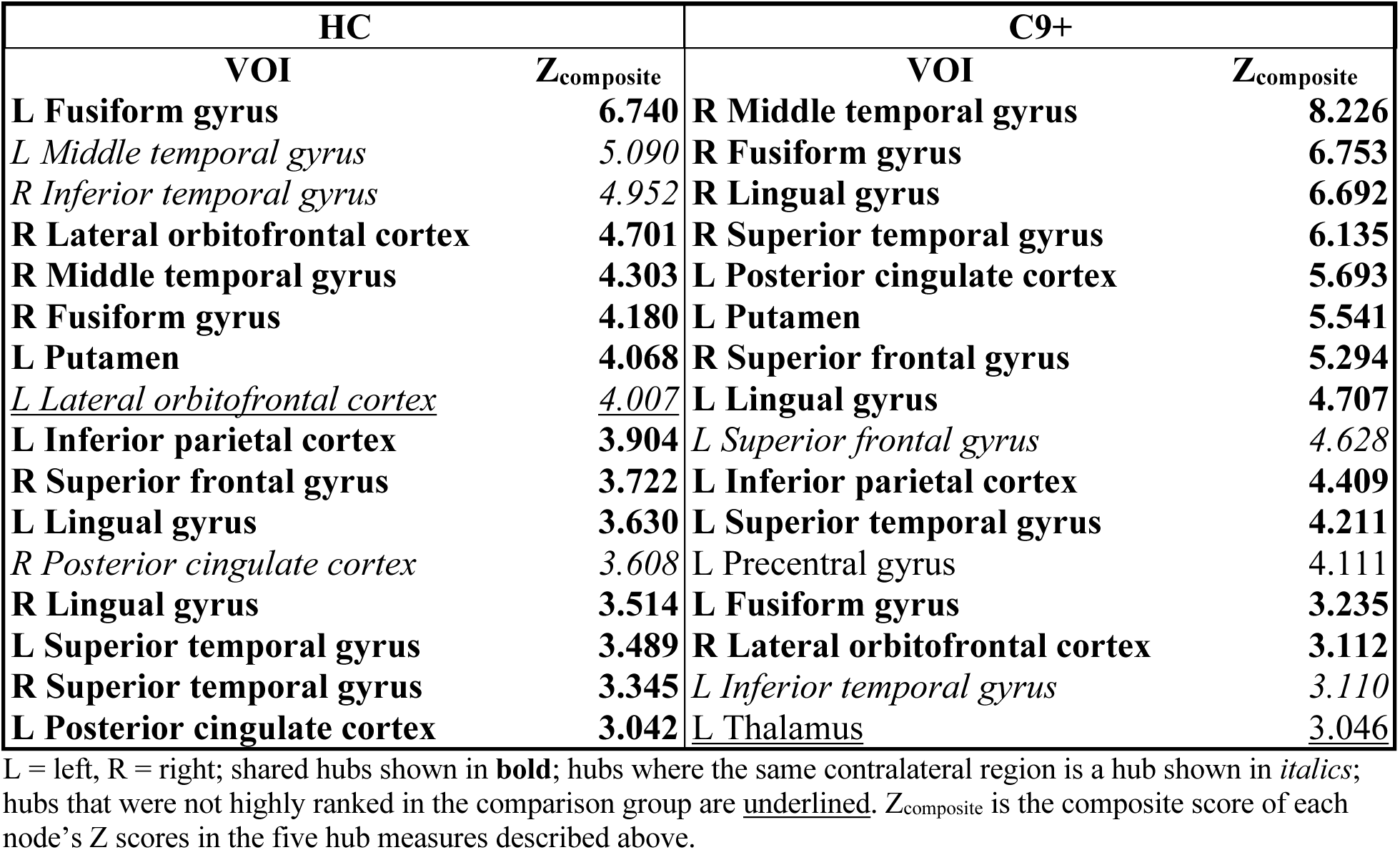
- Group hubs, defined as the top 20% of nodes based on a composite hub score (Z) for Healthy Controls and C9+ carrier groups.

| HC |  | C9+ |  |
| --- | --- | --- | --- |
| VOI | $Z_{\text{composite}}$ | VOI | $Z_{\text{composite}}$ |
| **L Fusiform gyrus** | **6.740** | **R Middle temporal gyrus** | **8.226** |
| *L Middle temporal gyrus* | *5.090* | **R Fusiform gyrus** | **6.753** |
| *R Inferior temporal gyrus* | *4.952* | **R Lingual gyrus** | **6.692** |
| **R Lateral orbitofrontal cortex** | **4.701** | **R Superior temporal gyrus** | **6.135** |
| **R Middle temporal gyrus** | **4.303** | **L Posterior cingulate cortex** | **5.693** |
| **R Fusiform gyrus** | **4.180** | **L Putamen** | **5.541** |
| **L Putamen** | **4.068** | **R Superior frontal gyrus** | **5.294** |
| <u>*L Lateral orbitofrontal cortex*</u> | <u>*4.007*</u> | **L Lingual gyrus** | **4.707** |
| **L Inferior parietal cortex** | **3.904** | *L Superior frontal gyrus* | *4.628* |
| **R Superior frontal gyrus** | **3.722** | **L Inferior parietal cortex** | **4.409** |
| **L Lingual gyrus** | **3.630** | **L Superior temporal gyrus** | **4.211** |
| *R Posterior cingulate cortex* | *3.608* | **L Precentral gyrus** | **4.111** |
| **R Lingual gyrus** | **3.514** | **L Fusiform gyrus** | **3.235** |
| **L Superior temporal gyrus** | **3.489** | **R Lateral orbitofrontal cortex** | **3.112** |
| **R Superior temporal gyrus** | **3.345** | *L Inferior temporal gyrus* | *3.110* |
| **L Posterior cingulate cortex** | **3.042** | <u>*L Thalamus*</u> | <u>*3.046*</u> |

L = left, R = right; shared hubs shown in **bold**; hubs where the same contralateral region is a hub shown in *italics*; hubs that were not highly ranked in the comparison group are underlined. $Z_{\text{composite}}$ is the composite score of each node's Z scores in the five hub measures described above.

#### 3.1.3 Edge analysis - C9+ vs. HC

**Figure 3** displays the edges that had significantly decreased functional connectivity in C9+ carriers compared to HC (p<0.001 uncorrected). There were no impaired intra-hemispheric connections in the left hemisphere. All connections with decreased functional connectivity were either inter-hemispheric or intra-hemispheric within the right hemisphere. More cortical connections were affected than subcortical connections. The right frontal lobe had the greatest number of reduced connections. Reduced connectivity was also seen for connections with the basal ganglia, temporal lobe, and parietal lobes. These edges connect regions involved in a broad range of cognitive functions. Additionally, several edges connecting motor-related regions were affected, including the right precentral and paracentral lobules. Other regions having multiple differing connections were the pars opercularis, pars triangularis, supramarginal gyrus, inferior temporal gyrus, and the left putamen.

**Figure 3.**
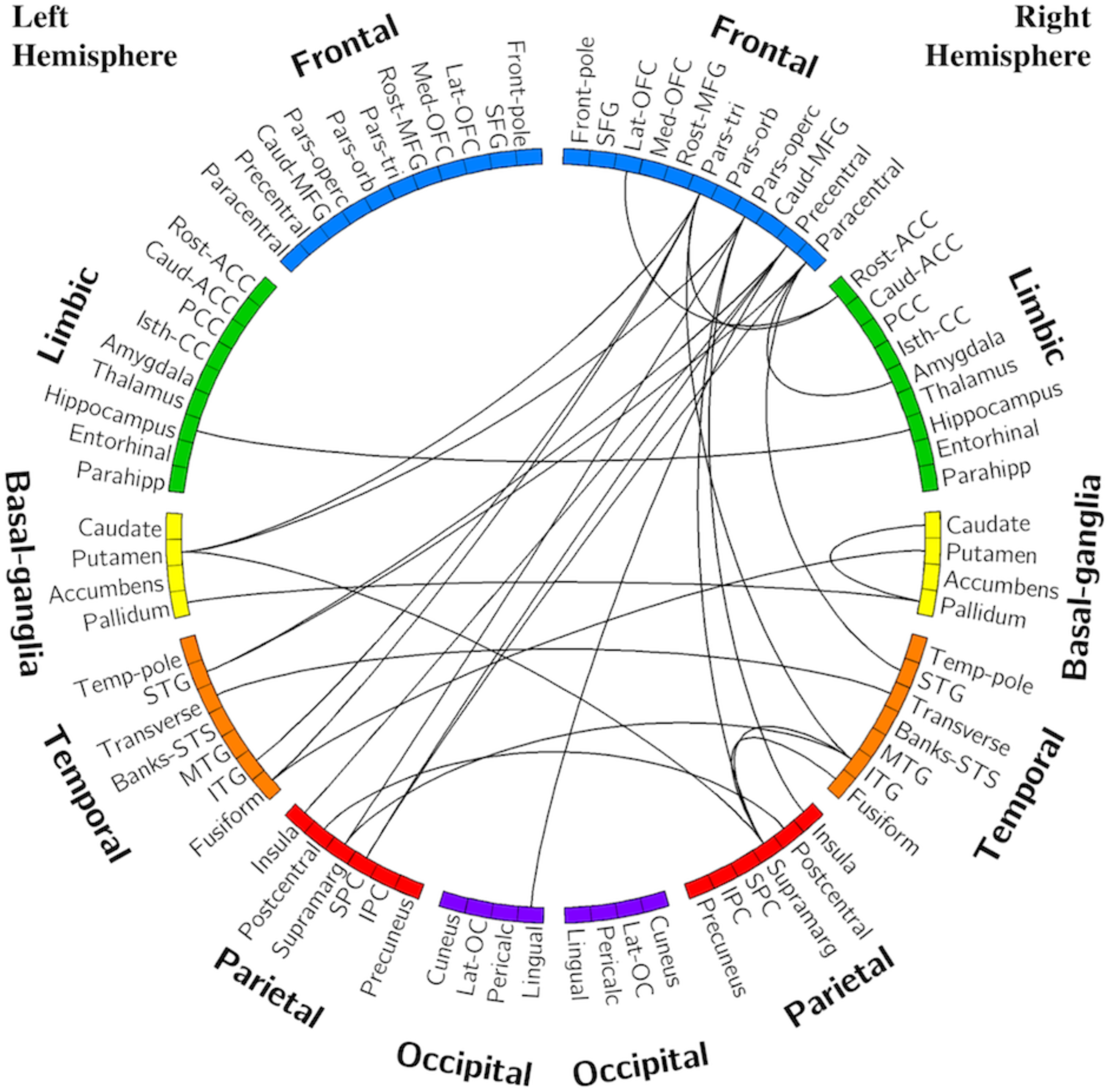
- Edges with reduced connectivity strength in C9+ carriers compared with HC. Significance was determined using permutation testing at p<0.001 uncorrected. See Table 2 for region name abbreviations.

### 3.2 Comparison of C9+ bvFTD/ALS-FTD carriers with C9+ ALS alone (bvFTD −)

#### 3.2.1 Global graph metrics – bvFTD+ vs. ALS-only

Within the symptomatic group of C9+ carriers, there were no significant differences in global metrics (network density, mean connection strength, clustering coefficient, path length, and modularity) between the 9 patients with ALS alone and the 9 patients with bvFTD or ALS-FTD.

#### 3.2.2 Network hubs - bvFTD+ vs. ALS-only

The ALS-only group had similar hubs to healthy controls, with the addition of the precentral gyri and right caudate. In contrast, the bvFTD+ group had several nodes with hub scores that were ranked much lower than in the ALS-only group and healthy controls. These hubs included bilateral thalamus, right hippocampus, and right lateral occipital cortex. Hubs are listed in **Table 5**.

**Table 5.** **-** Group hubs, defined as top 20% of nodes based on composite hub score for symptomatic patients with and without bvFTD.

| C9+ bvFTD+ (n=9) |  | C9+ ALS-only (n=9) |  |
| --- | --- | --- | --- |
| VOI | Z<sub>composite</sub> | VOI | Z<sub>composite</sub> |
| L Posterior cingulate cortex | 9.512 | **R Superior temporal gyrus** | **8.346** |
| **L Superior temporal gyrus** | **7.121** | **R Lingual gyrus** | **7.159** |
| **R Superior frontal gyrus** | **6.652** | **R Middle temporal gyrus** | **6.744** |
| **L Superior frontal gyrus** | **6.437** | *L Middle temporal gyrus* | *6.483* |
| L Putamen | 6.391 | **L Superior temporal gyrus** | **5.412** |
| **R Superior temporal gyrus** | **5.528** | R Caudate | 5.135 |
| R Fusiform gyrus | 5.102 | R Banks STS | <u>4.803</u> |
| <u>R Thalamus</u> | <u>5.022</u> | **R Inferior parietal cortex** | **4.639** |
| <u>L Thalamus</u> | <u>4.732</u> | **R Superior frontal gyrus** | **4.535** |
| **R Lingual gyrus** | **4.629** | <u>R Precentral gyrus</u> | <u>4.425</u> |
| <u>R Lateral occipital cortex</u> | <u>4.124</u> | L Lingual gyrus | 4.330 |
| <u>L Pericalcarine cortex</u> | <u>4.069</u> | <u>R Caudal anterior cingulate cortex</u> | <u>3.925</u> |
| *L Inferior temporal gyrus* | 3.797 | **L Superior frontal gyrus** | **3.862** |
| **R Middle temporal gyrus** | **3.732** | <u>R Isthmus of the cingulate cortex</u> | <u>3.810</u> |
| **R Inferior parietal cortex** | **3.728** | *R Inferior temporal gyrus* | *3.734* |
| <u>R Hippocampus</u> | <u>3.626</u> | <u>L Precentral gyrus</u> | <u>3.596</u> |

L = left, R = right; shared hubs shown in **bold**; hubs where the same contralateral region is a hub shown in *italics*; hubs that were not highly ranked in the comparison group are underlined. Z<sub>composite</sub> is the composite score of each node's Z scores in the five hub measures described above.

The correlations between the hub score and the FBI and ALSFRS-R scores were computed for each of the unique hubs within each group. Three of the unique hubs in bvFTD+ patients correlated with FBI scores across all symptomatic C9+, indicating that behavioral impairment was associated with higher node strength (**Figure 4**). These hub nodes were the left thalamus (R=0.443, p=0.066), right thalamus (R=0.471, p=0.049), and right hippocampus (R=0.525, p=0.025).

**Figure 4.**
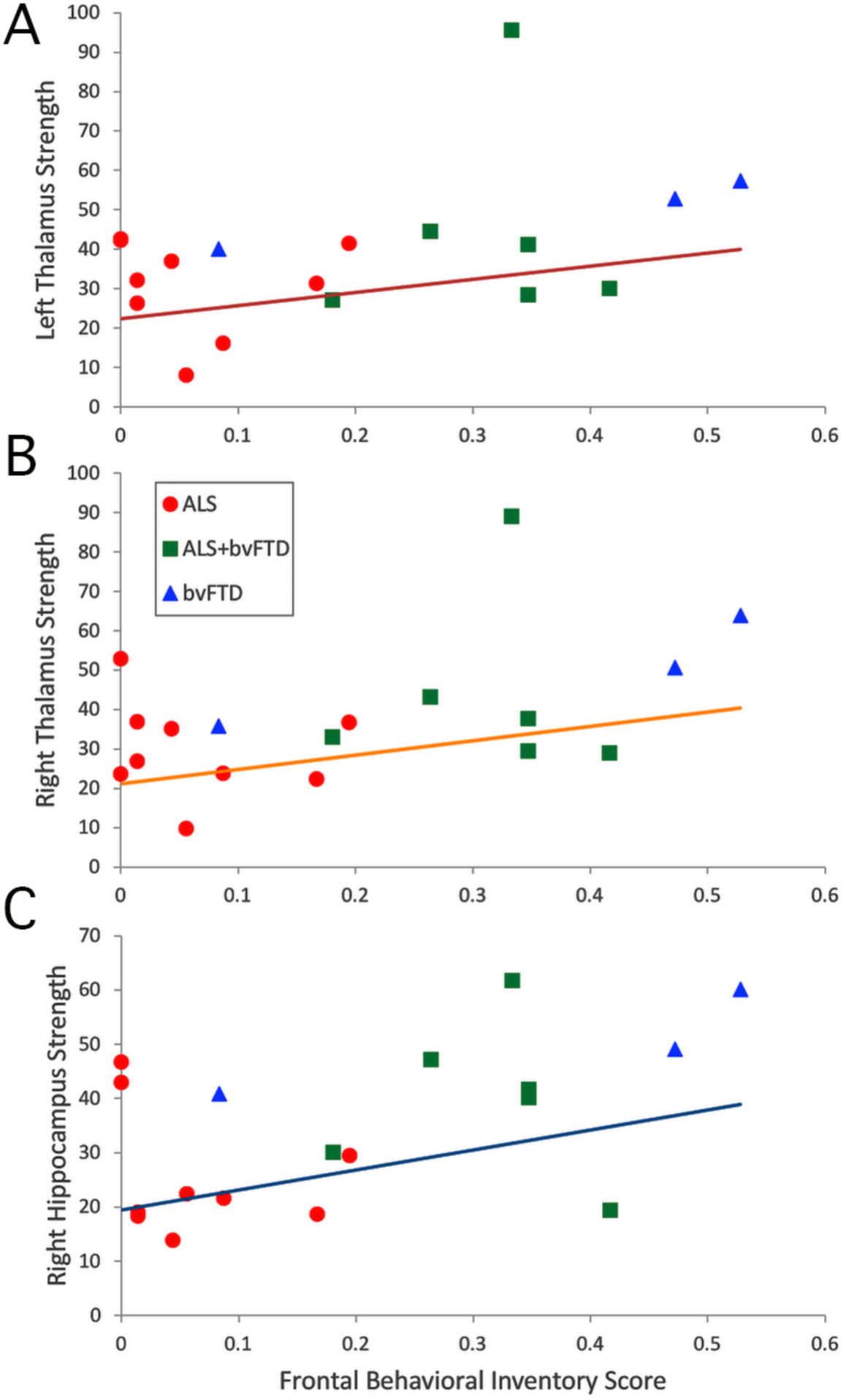
– Correlation across all symptomatic C9+ carriers of Frontal Behavioral Inventory Scores with node strength of hubs that were more highly ranked in the bvFTD+ group than ALS-only group. (A) Left thalamus, R=0.443, p=0.066 (B) Right thalamus, R=0.471, p=0.049 (C) Right hippocampus, R=0.525, p=0.025.

#### 3.2.3 Edge analysis - C9+ bvFTD+ vs. ALS-only

Several edges had reduced connectivity strength in the ALS-only group compared to the bvFTD+ group. All statistically significant differences consisted of decreases in connection strength in the ALS-only group compared with the bvFTD+ group. The frontal and temporal regions exhibited the greatest number of decreased connections (**Figure 5**).

**Figure 5.**
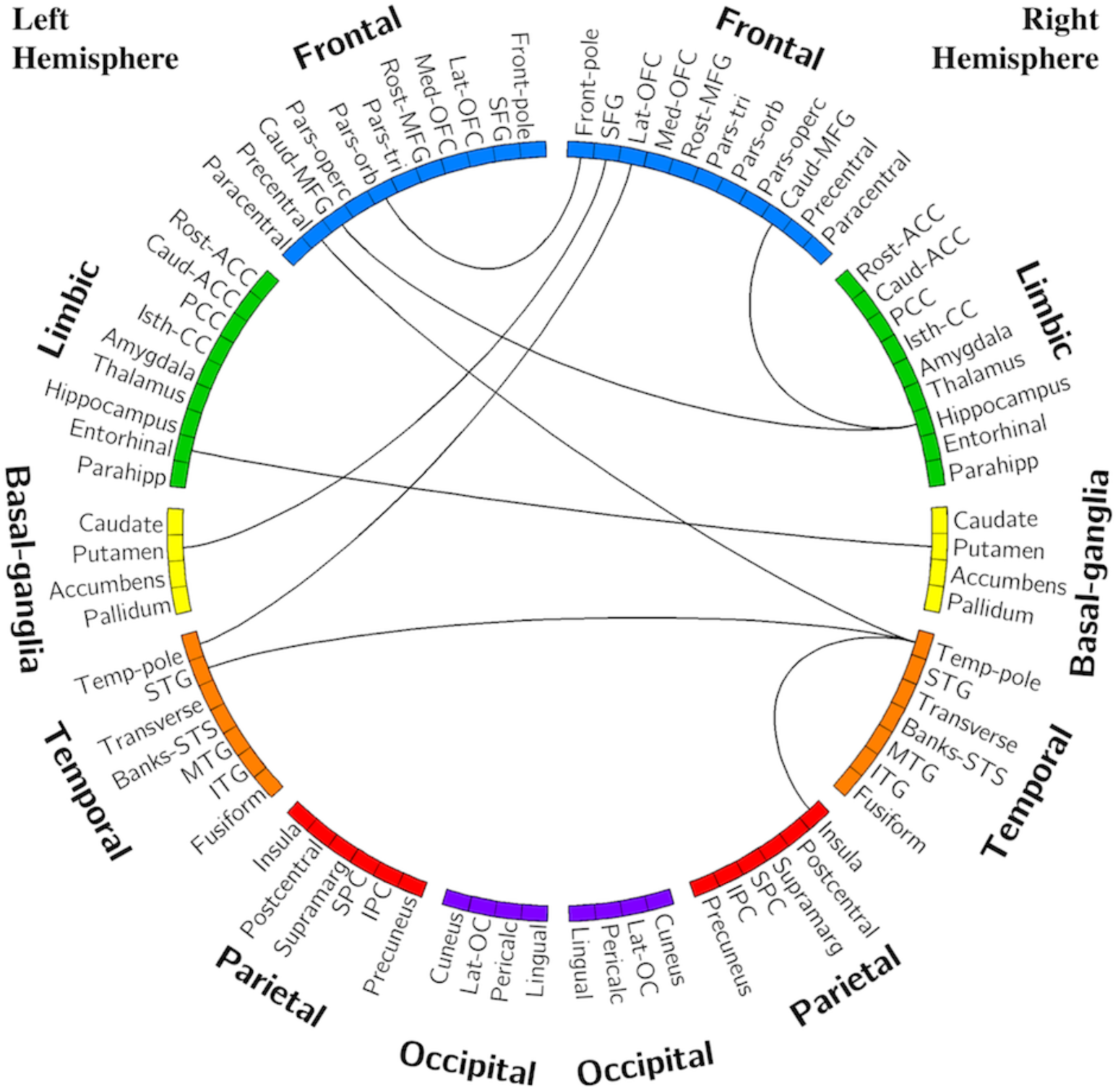
- Edges with reduced connectivity strength in ALS-only group compared with bvFTD+/ALS-FTD. Significance was determined using permutation testing at p<0.001 uncorrected. See Table 2 for region name abbreviations.

## 4 Discussion

In this study, we used graph theory metrics to explore alterations in network organization in *C9orf72* mutation carriers and network correlations with motor and cognitive-behavioral functional measures. Graph theory allows exploration of network organization and functionality as a whole, rather than correlations between activity in discrete regions. We found that global network measures of connectivity were reduced in a cohort of C9+ carriers with heterogenous symptoms compared to healthy controls. Because *C9orf72* expansion mutations cause motor and cognitive-behavioral symptoms across the ALS-FTD spectrum, we had anticipated that brain regions known to be affected in sporadic forms of both diseases would exhibit connectivity changes. This was mostly confirmed, with reduced connectivity specifically found in connections involving the frontal and temporal regions and the right motor cortex, regions known to be involved in cognitive and motor function. These findings are consistent with many prior studies in sporadic and C9+ FTD (13, 15, 22, 33, 46-52). However, as previously noted, the literature on resting state connectivity in sporadic ALS has many discordant results. Our findings are consistent with studies showing decreased connectivity in ALS (29, 30, 32, 51, 53). The nodes with the greatest numbers of impaired connections in the C9+ population represented regions that are involved in networks previously described as being affected in ALS and bvFTD. These regions–the right precentral, paracentral, supramarginal, and inferior temporal gyri, the pars opercularis, pars triangularis, and the left putamen–are parts of the sensorimotor, salience, and central executive networks (54, 55).

Interestingly, while overall connectivity was decreased in the C9+ group, there appeared to be no substantial reorganization of network hubs. This indicates a relatively diffuse global decline in connectivity. Further underscoring this point, the edge analysis found no regions with increased connectivity in the C9+ network compared to HCs. This relative preservation of network organization may be interpreted as supporting the proposal that networks can compensate for low grade degeneration for a substantial period in order to maintain clinical function (56, 57). An alternative possibility is that pooling C9+ ALS, ALS-FTD, bvFTD, and presymptomatic carriers into one analysis group masked phenotype-specific network alterations.

To explore this possibility, we compared subgroups of C9+ symptomatic patients with and without FTD. This analysis found no significant differences in global network measures between subgroups, indicating that the global decrease in connectivity observed in the full C9+ cohort was likely not solely driven by a large change in one of the phenotypic subgroups. However, nodal graph theory metrics revealed some organizational differences between subgroups. Both of the symptomatic phenotypic subgroups had several hubs that that were unique in comparison with the healthy controls and from the other subgroup. In the ALS-only group (i.e., patients without FTD), the bilateral precentral gyri emerged as group-specific hubs. This implies that the ALS-only subgroup either had small increases in motor cortex connectivity or that the motor cortex connectivity remained relatively stable in the face of declining connectivity of the more active nodes. This phenomenon is consistent with resting state studies of patients with sporadic ALS that demonstrate localized increases in connectivity of motor regions (15, 25, 58, 59), and could represent a compensatory mechanism or the relative resilience of the motor network.

The bvFTD+ group had a greater number of subcortical hubs. The bilateral thalami and the right hippocampus were hub nodes in the bvFTD+ group and were not highly ranked in the ALS-only group or in healthy controls. The greater dependence on subcortical nodes could arise as a consequence of structural changes, including cortical atrophy (10, 15, 60, 61). The emergence of the thalamus as an important hub in C9+ bvFTD was somewhat surprising given that thalamic atrophy occurs in bvFTD (13, 16, 62) and in C9+ patients (8, 10, 15, 21). It also seems to conflict with resting state studies of bvFTD that report decreases in thalamic connectivity (47, 63). This may reflect differences between seed-based analysis methods (which may neglect global neuromodulatory changes) versus the whole-brain approach used here. On balance, these modulated connections could result in the thalamus having a more prominent role as a network hub. The emergence of the hippocampus as a hub may account for the preservation of memory in the first few years of bvFTD symptoms (39, 62), although there are a few reports of hippocampal atrophy in bvFTD (48, 64).

To investigate the relationship between disease severity and network hub changes, we evaluated the correlation between hub scores of hubs unique to each subgroup and the FBI and ALSFRS-R scores of symptomatic C9+ carriers. The hub scores of the bilateral thalami and right hippocampus correlated with behavioral impairment as measured by the FBI. There was also significantly higher connectivity between the right hippocampus and the right and left middle frontal gyrus in the analysis of individual connections in the bvFTD+ group. Therefore, as the thalamus and hippocampus became more hub-like, patients exhibited more severe behavioral impairment. In contrast, there was no association between the hub scores and the ALSFRS-R scores for the unique hubs in the ALS-only group. The connectivity of these motor regions appears to change independently from this measure of motor symptom severity.

There are limitations in the present study that warrant discussion. First, because no subjects with sporadic disease were considered for this study, it is impossible to determine if any effects described here are unique to familial (and specifically C9-linked) disease. Second, as a result of our relatively small sample size, there is a large amount of heterogeneity in the study population and functional connectivity data. Subjects were grouped according to meeting diagnostic criteria for ALS and/or bvFTD; however, minor symptoms that were insufficient to meet criteria for a clinical diagnosis could nevertheless affect neural activity. Future studies with larger subgroups are warranted. Third, due to the wide variation in disease severity and duration amongst subjects, our cross-sectional design may not capture the full extent of functional connectivity changes in C9+ disease; a longitudinal study would be better suited to explore the evolution of functional connectivity changes across the ALS-FTD spectrum over time.

## 5 Conclusion

Carriers of the *C9orf72* repeat expansion mutation have decreased functional connectivity at rest compared with healthy controls. Global network organization is generally preserved, although local network alterations emerge in C9+ carriers with ALS versus FTD. Subcortical regions, including the bilateral thalami and the right hippocampus, emerge as hubs associated with the severity of behavioral impairment.

## Data Availability

The corresponding author will make de-identified data available upon reasonable request.

## Author Contributions

RS contributed to data collection, analytical design, data analysis and interpretation, and manuscript writing. MC contributed to data collection, data interpretation, and manuscript editing. MKF contributed to project conception and design, data collection, data interpretation, and manuscript drafting.

## Acknowledgements

Recruitment was made possible in part by ATSDR’s National ALS Registry Research Notification Mechanism

(https://www.cdc.gov/als/ALSResearchNotificationClinicalTrialsStudies.html).

We gratefully acknowledge Jennifer Farren, R.N. for patient coordination, the neurologists who referred patients, and the patients and caregivers whose participation was invaluable. We also give our thanks to Laura Braun and Laura Danielian for their contributions in data collection. This work utilized the computational resources of the NIH HPC Biowulf cluster (https://hpc.nih.gov).

